# Use of tryptic peptide MALDI mass spectrometry imaging to identify the spatial proteomic landscape of colorectal cancer liver metastases

**DOI:** 10.1101/2024.01.24.24301748

**Authors:** Celine Man Ying Li, Matthew T Briggs, Yea-Rin Lee, Teresa Tin, Clifford Young, John Pierides, Gurjeet Kaur, Paul Drew, Guy J Maddern, Peter Hoffmann, Manuela Klingler-Hoffmann, Kevin Fenix

## Abstract

Colorectal cancer (CRC) is the second leading cause of cancer-related deaths worldwide. CRC liver metastases (CRLM) are often resistant to conventional treatments, with high rates of recurrence. Therefore, it is crucial to identify biomarkers for CRLM patients that predict cancer progression. This study utilised matrix-assisted laser desorption/ionisation mass spectrometry imaging (MALDI-MSI) in combination with liquid chromatography-tandem mass spectrometry (LC-MS/MS) to spatially map the CRLM tumour proteome. CRLM tissue microarrays (TMAs) of 84 patients were analysed using tryptic peptide MALDI-MSI to spatially monitor peptide abundances across CRLM tissues. Abundance of peptides was compared between tumour vs stroma, male vs female and across three groups of patients based on overall survival (0-3 years, 4-6 years, and 7+ years). Peptides were then characterised and matched using LC-MS/MS. A total of 471 potential peptides were identified by MALDI-MSI. Our results show that two unidentified *m/z* values (1589.876 and 1092.727) had significantly higher intensities in tumours compared to stroma. Ten *m/z* values were identified to have correlation with biological sex. Survival analysis identified three peptides (Histone H4, Hemoglobin subunit alpha, and Inosine-5’-monophosphate dehydrogenase 2) and two unidentified *m/z* values (1305.840 and 1661.060) that were significantly higher in patients with shorter survival (0-3 years relative to 4-6 years and 7+ years). This is the first study using MALDI-MSI, combined with LC-MS/MS, on a large cohort of CRLM patients to identify the spatial proteome in this malignancy. Further, we identify several protein candidates that may be suitable for drug targeting or for future prognostic biomarker development.

## Introduction

Colorectal cancer (CRC) is the third most common cancer and the second leading cause of cancer-related deaths worldwide [1]. Approximately 50% of CRC cases metastasise to the liver [2]. CRC liver metastasis (CRLM) is normally treated with surgical resection and chemotherapy. However, most CRLM cases present with multiple liver metastases or have patients with poor health conditions that prevents surgery [3]. Most CRLMs ultimately develop chemo-resistance [4]. Even with curative intent, CRLM has a relapse rate of 50% [5]. Together these factors contribute to CRLM having a 5-year survival rate of 30% [2]. Currently, there are no predictive biomarkers for CRLM recurrence or survival. CRLM surveillance currently involves frequent computed tomographic (CT) scans and monitoring for blood carcinoembryonic antigen (CEA) levels [6, 7]. However, these surveillance strategies have failed to detect early relapse, contributing to disease progression. Thus, there is an urgent need for predictive markers for CRLM survival [5].

Quantitative image analysis for tissue histological biomarkers based on routine histochemistry or immunohistochemistry has been used for clinical diagnostics and biomarker research [8]. In cancer research, recent advances in mass spectrometry have made proteomics a useful approach for biomarker discovery [9, 10]. In particular, matrix-assisted laser desorption/ionisation (MALDI) mass spectrometry imaging (MSI) generates unbiased spatial intensity maps of molecules such as intact proteins, tryptic peptides, *N-*glycans, lipids or metabolites, which allows for the spatial mapping of tumour tissues. MALDI-MSI has been demonstrated to predict metastatic potential, disease recurrence, and survival in variety of primary cancers [11–13].

In the context of CRC, MALDI-MSI has been extensively applied to primary tumours and has identified prognostic indicators for overall survival [14–16]. For example, multiple analytes have been shown to be associated with clinical outcomes such as tumour stage, grade, metastasis, and cancer cell proliferation in primary CRC. However, there are a limited number of MALDI-MSI studies on CRLM. To the best of our knowledge, only two reports have applied MALDI-MSI to detect lipid [17] and phospholipid [18] signatures on CRLM tissue. These studies identified tumour localised lipid signatures that could have prognostic potential or can be therapeutic targets. Whereas two additional MALDI-MSI studies identified the spatial proteome with limited sample sizes preventing any correlation with survival [19, 20].

In this study, we applied tryptic peptide MALDI-MSI on a cohort of Australian CRLM patients and identified peptides that discriminate between tumour features, patient characteristics, and survival outcomes. Subsequently, liquid chromatography-tandem mass spectrometry (LC-MS/MS) was performed on tryptic peptides obtained from consecutive tissue sections to identify the associated proteins [21]. We identified discriminative proteins related to tumour features, and potential prognostic biomarkers that were associated with worse clinical outcomes in CRLM patients.

## Material and Methods

### Sample Collection and Tissue Specimens

A retrospective cohort of CRC patients with liver metastases were identified using the South Australian Metastatic Colorectal Cancer Registry (SAmCRC) [22]. From that cohort, patients who had undergone liver resection were selected for the study. Their corresponding FFPE tissue blocks were retrieved from SA Pathology sites based at The Queen Elizabeth Hospital, Royal Adelaide Hospital and Flinders Medical Centre, Adelaide, South Australia. Retrieved cases were reviewed by a clinical pathologist (J.P) for study suitability. Demographics and clinicopathological characteristics of patients were shown in Table 1. This study was approved by the Human Research Ethics Committee of the Central Adelaide Local Health Network under protocol number 12237.

**Table 1.**
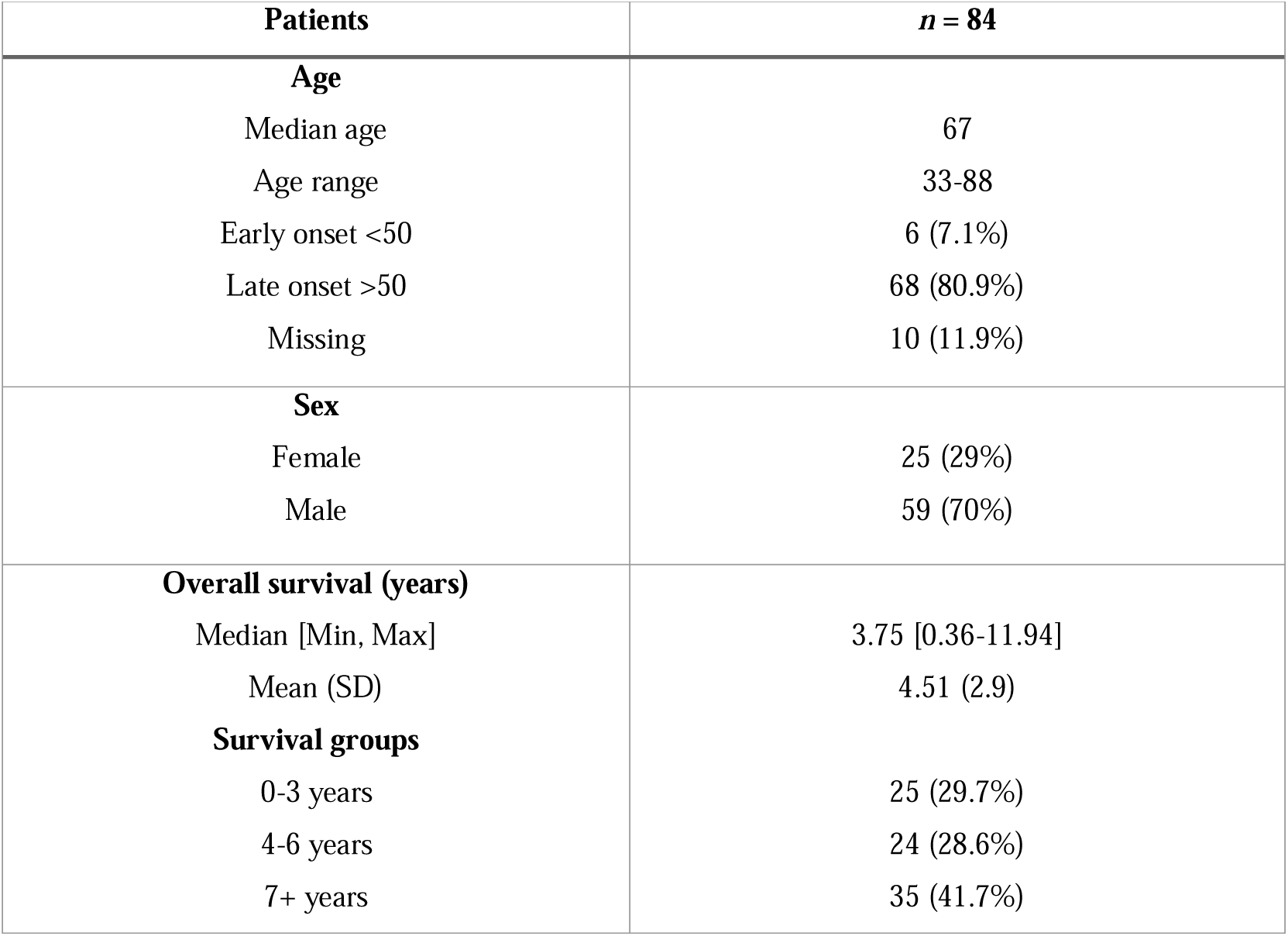
Demographic and clinicopathological characteristics of Australian CRLM patient cohort (n=84).

| **Patients** | ***n* = 84** |
| --- | --- |
| **Age** |  |
| Median age | 67 |
| Age range | 33-88 |
| Early onset <50 | 6 (7.1%) |
| Late onset >50 | 68 (80.9%) |
| Missing | 10 (11.9%) |
| **Sex** |  |
| Female | 25 (29%) |
| Male | 59 (70%) |
| **Overall survival (years)** |  |
| Median [Min, Max] | 3.75 [0.36-11.94] |
| Mean (SD) | 4.51 (2.9) |
| **Survival groups** |  |
| 0-3 years | 25 (29.7%) |
| 4-6 years | 24 (28.6%) |
| 7+ years | 35 (41.7%) |

### Tissue Microarray Construction

Previously selected tissue blocks were annotated by a pathologist (J.P), clearly distinguishing regions with viable tumours from normal liver. Two tumour cores and one normal liver core per patient were inserted into the TMA recipient block. Four TMA blocks were analysed, totalling 168 CRLM tumour cores from 84 patients. TMAs were constructed using a TMA Master II (3DHISTECH, Budapest, Hungary). The completed block was sealed with paraffin and stored at 4°C.

### Hematoxylin and Eosin (H&E) Staining

FFPE tissue blocks were cut by microtome into 6 µm sections. They were first deparaffinized and rehydrated with xylene for 3 x 5 mins, dipped in ethanol (100%) for 3 x 4 mins, ethanol (90%, *v/v*) for 2 mins, ethanol (70%, *v/v*) for 2 mins, and washed with distilled water for 1 min. Subsequently, slides were stained with hematoxylin for 5 mins, then washed with running tap water for 5 mins. Slides were then dipped into 0.3% hydrochloric acid with ethanol (75%, *v/v*) for 10 seconds, followed by washing with tap water for 10 mins. Tissues were stained with eosin for 1 min and washed with ethanol (100%) for 1 min. The H&E-stained TMA slides were scanned by a NanoZoomer 1 Digital Slide Scanner (Hamamatsu Photonics K.K, Japan). Images were acquired and viewed using NDP.view2 software (Hamamatsu).

### Tryptic Peptide MALDI-MSI Sample Preparation

Tryptic peptide MALDI-MSI was performed on the sectioned TMAs following the protocol as previously described, with minor modifications [23]. Briefly, 6 µm sectioned TMAs on indium tin oxide (ITO) slides were washed with RCL premium grade xylene (100%) for 2 x 5 min, HPLC grade ethanol (100%) for 2 x 2 min, and 10 mM ammonium bicarbonate for 2 x 5 min. Slides were then boiled with 10 mM citric acid (pH 6) in a conventional microwave (1250 W, Model MS2540SR, LG, China), followed by pulse-heating for 10 min and heating at 98 °C on a heating block for 30 min. Slides were cooled down at room temperature prior to washing with 10 mM ammonium bicarbonate for 2 x 1 min. Subsequently, slides were dried at ambient conditions, marked with a water-based white-out, and scanned using a flattop scanner (Epson Perfection V600 Photo, Model J25A, Epson, Indonesia) to teach the instrument during MALDI-MSI data acquisition.

Next, 20 µg of trypsin gold (Promega) in 200 µL of 25 mM ammonium bicarbonate and acetonitrile (10%, *v/v*) was deposited onto slides using an ImagePrep instrument (Bruker Daltonic, Germany) in 30 cycles with a fix nebulization time of 1.2 sec and drying time of 15 sec followed by incubation at 37 °C for 2 h in a humidified chamber containing potassium sulphate (53.3 g) and MilliQ water (18.2 g). Next, 7 mg/mL of alpha-cyano-4-hydroxycinnamic acid (HCCA) in acetonitrile (50%, *v/v*) and trifluoroacetic acid (TFA, 0.2%, *v/v*) was deposited onto slides using an iMatrix Spray instrument (Tardo, Switzerland). Instrument-specific settings were as follows: 60 mm height, 1 mm line distance, 180 mm/s speed, 1 µL/cm^2^ density, 30 cycles and 15 sec delays.

### MALDI-MSI Data Acquisition

MALDI-MSI data was acquired on a timsTOF fleX MS instrument (Bruker Daltonic, Germany), controlled by timsControl (v3.0, Bruker Daltonics) and flexImaging (v7.0, Bruker Daltonics) in positive mode in the mass range of *m/z* 700-3200. The instrument was calibrated using an internal calibrant [24]. Instrument-specific settings were as follows: a transfer time of 180 µs, a pre-pulse storage of 25 µs, a collision RF of 4000 Vpp, a collision energy of 10 eV, a funnel 1 (accumulation) RF of 500 Vpp, a funnel 2 (analysis) RF of 500 Vpp, a multipole RF of 500 Vpp and a quadrupole ion energy of 5 eV. Laser power was set at 60% and pixel resolution at 50µm.

### MALDI-MSI Data Analysis

MALDI-MSI data was imported into SCiLS Lab v2023a (Bruker Daltonics, Germany) and pre-processed by TopHat baseline subtraction and root mean square (RMS) normalisation. Individual TMA cores were annotated based on pathological annotations by a pathologist (G.K.) from consecutive H&E-stained images scanned with NanoZoomer (Hamamatsu). The feature finding tools were then used to create a list of putative peptides. The feature finding uses the T-Rex^2^ algorithm, and very strong filtering with 5% coverage and 0.5% relative intensity threshold were applied. For comparisons of peak intensities between the groups, the RMS normalised average spectra of each mass was exported as Excel files and statistical analysis performed using GraphPad Prism (v 9.0, San Diego, CA).

### LC-MS/MS Sample Preparation

CRLM TMAs were sectioned using a microtome (Microm HM 325) at 6 µm thickness and placed directly into 1.5 mL Eppendorf Protein LoBind Tubes. Paraffin was removed by the addition of 100 µL of xylene (100%) for 2 min and 100 µL of ethanol (100%) for 2 min. Sections were then rehydrated by the addition of 100 µL of Milli-Q water (100%) for 2 x 2 min and 100 µL of 50 mM ammonium bicarbonate (100%) for 2 x 2 min. Then the tissue was boiled in 30 µL of 10 mM citric acid at pH 6 at 98 °C for 20 min in a Thermomixer (1500 rpm), followed by 80 °C for 2 h. Tubes were cooled to room temperature and then centrifuged at 20,000 x g and 4 °C for 30 min. Proteins were denatured from the tissue by adding 30 µL of 8 M urea, followed by incubation at room temperature for 30 min. Subsequently, samples were analysed using a modified tryptophan assay to determine protein concentrations [25].

Protein reduction was by incubation with 2.5 µL of 0.2 M dithiothreitol (DTT) at room temperature for 1 hr on a Thermomixer (500 rpm). Proteins were then alkylated by the addition of 2.8 µL of 0.275 M 2-chloroacetamide (CA) and incubation in the dark for 30 min on a shaking Thermomixer. Protein digestion was performed with sequencing grade trypsin (Promega, USA) (1:50) at 37 °C overnight on a shaking Thermomixer, with digestion stopped by the addition of 10% formic acid (5 µL). Finally, tryptic peptides were desalted using C18 ZipTips (Millipore, Ireland) according to the manufacturer’s instructions.

### LC-MS/MS Data Acquisition

LC-MS/MS analysis was conducted on an EASY-nLC 1200 system coupled to an Orbitrap Exploris 480 mass spectrometer (Thermo Scientific, Bremen, Germany). Peptides (approximately 500 ng) were resuspended in 0.1% formic acid and loaded onto a 25 cm fused silica column (75 µm inner diameter, 360 µm outer diameter) heated to 50 ^°^C. The column was packed with 1.9 µm ReproSil-Pur 120 C18-AQ particles (Dr. Maisch, Ammerbuch, Germany). Peptides were separated over a 30 min linear gradient (3 to 24% acetonitrile in 0.1% formic acid) at a flow rate of 300 nL/min. A compensation voltage of –45 V was applied from a FAIMS Pro interface (Thermo Scientific) to minimise the impact of interfering ions before implementing a data-independent acquisition (DIA) MS method. Briefly, an MS scan (*m/z* 395 to 905) was acquired at resolution 30000 (*m/z* 200) in positive ion mode before isolated precursors (50 x 10 *m/z* windows, 1 *m/z* overlap) were fragmented with higher energy collision dissociation (27.5% normalised collision energy). The resulting MS/MS spectra (*m/z* 145 to 1450) were measured at resolution 22500.

### LC-MS/MS Data Analysis

Raw data files were processed with Spectronaut v17.4.230317 (Biognosys AG, Schlieren, Switzerland). A direct DIA+ analysis was conducted by searching a UniProt human proteome database (Release 2022_04, 20607 canonical entries) with BGS factory settings. We manually matched single charged *m/z* values obtained from the MALDI-MSI data with singly charged *m/z* values calculated from doubly or triply charged peptides identified using Spectronaut analysis peptides with a peptide tolerance of +/− 16 ppm.

## Results

### Identification of tryptic peptide signatures in CRLM samples using MALDI-MSI

To identify tryptic peptides associated with CRLM, we first constructed four TMAs from 84 CRLM patients consisting of 168 CRLM tumour tissue cores (two per patient, Table 1). The TMAs were sectioned onto conductive ITO slides and prepared for MALDI-MSI analysis. A comprehensive and detailed MALDI-MSI workflow is presented in Fig. 1A. For the analysis of the processed MALDI-MSI data, tumour cores were annotated as tumour (cancer cell rich) and intratumoral stroma (cancer cell poor/free) regions in SCiLS Lab by matching pathologist annotated H&E regions overlaid with the MALDI-MSI optical image (Fig. 2). Subsequently, the feature finding tool in SCiLS Lab was used to generate a list of 471 putative peptides by applying very strong filtering with 5% coverage and 0.5% relative intensity threshold. Next, the receiver operating curve (ROC) discriminating feature tool was used to identify *m/z* values with the highest area under the curve (AUC) when comparing between tumour and stroma, male and female patients and across three groups of patients’ overall survival (0-3 years, 4-6 years, and 7+ years). Furthermore, denatured proteins from consecutive tissue sections were then digested in-solution into tryptic peptides for LC-MS/MS analysis to identify proteins by matching peptide sequences with measured *m/z* values by MALDI-MSI (Fig. 1B). To achieve an extensive list of peptides identified by LC-MS/MS, the raw data was analysed in Spectronaut and resulted in the identification of 3332 proteins from CRLM samples.

**Figure 1.**
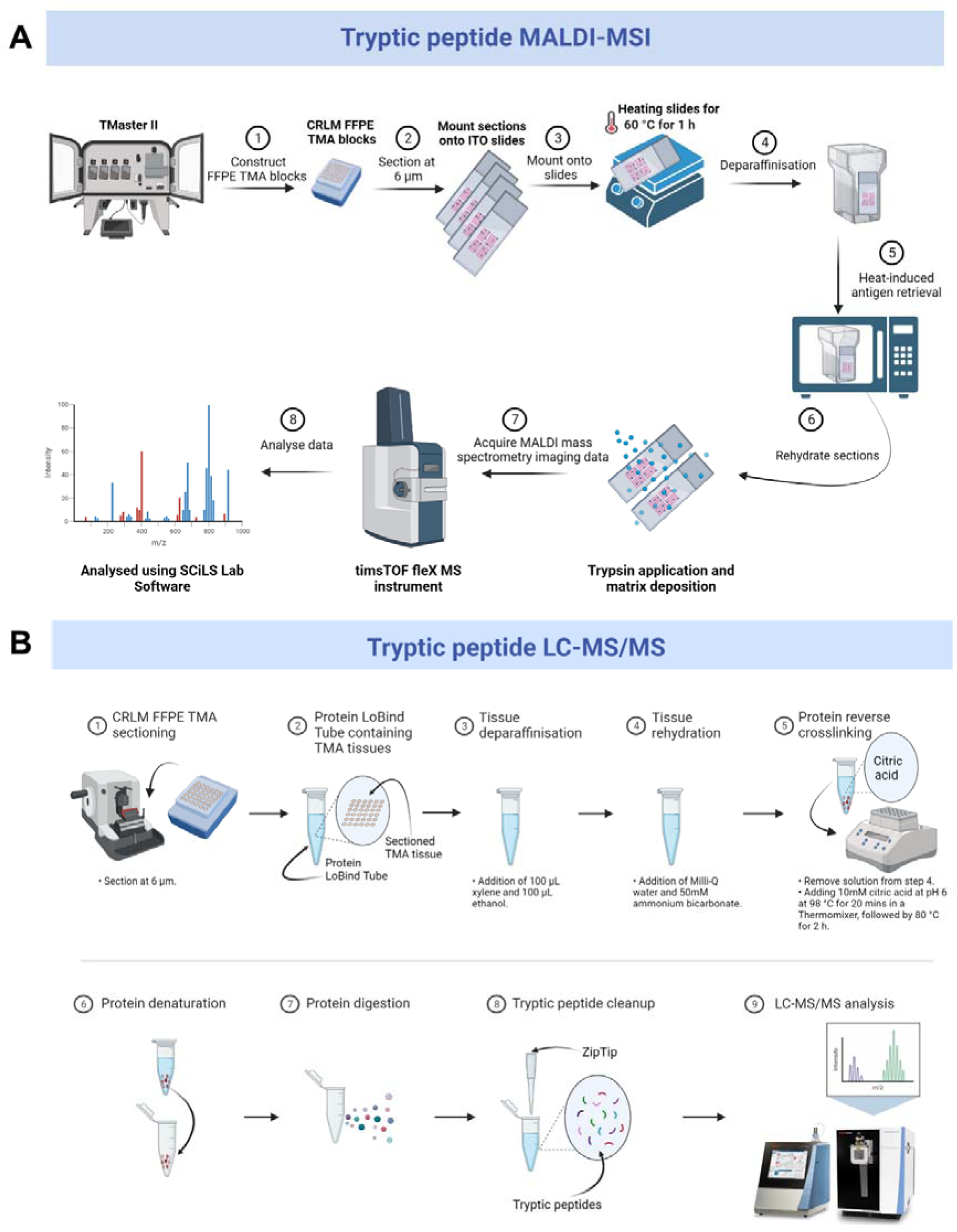
Overview of (A) MALDI-MSI and (B) LC-MS/MS workflows for spatial mapping and characterisation of tryptic peptides from CRLM FFPE tissues (n=84). Created with BioRender.com.

**Figure 2.**
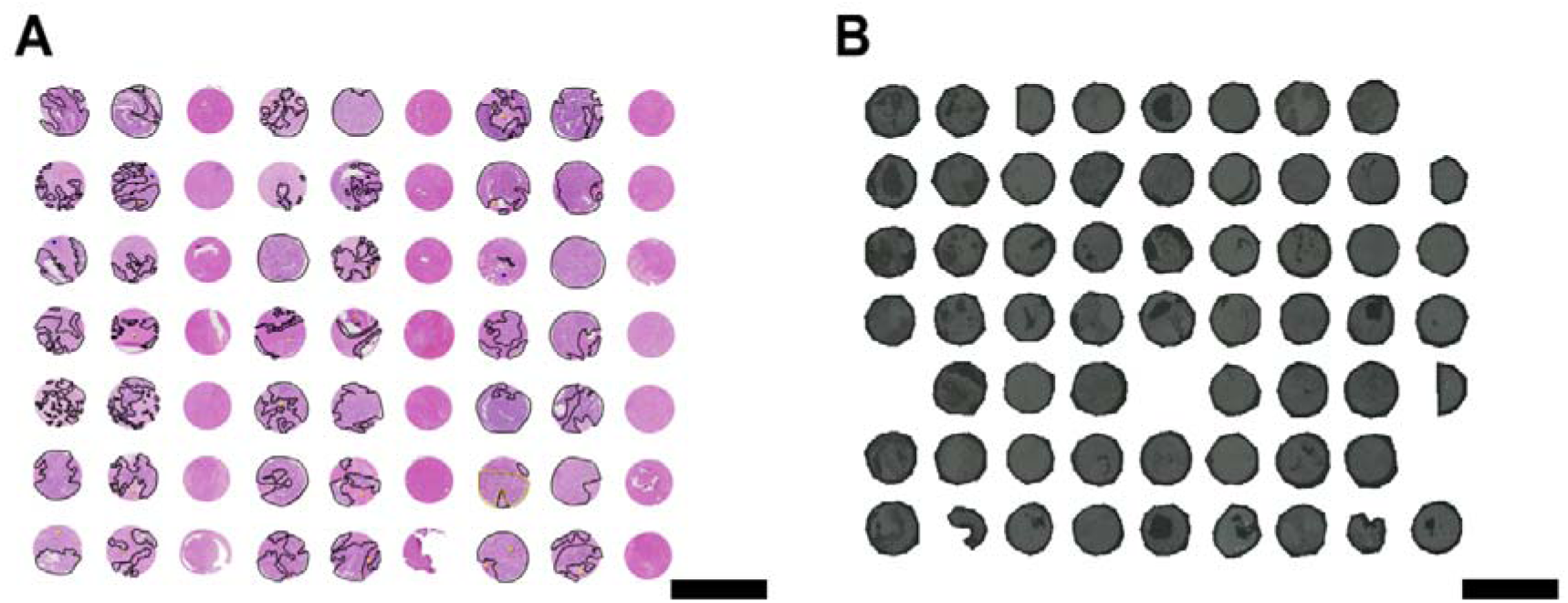
Representative scanned images of (A) H&E CRLM TMA cores, and (B) optical CLRM TMA cores. CRLM tumour regions and CRC tumour regions (control) in (A) were annotated by a pathologist, as shown in black and green, respectively. Scale bars (black lines) represent 3mm.

### Identification of tryptic peptides associated with CRLM tumour regions

First, to identify tryptic peptides that significantly distinguished between tumour and stroma, we selected MALDI-MSI *m/z* values with the highest AUC values to discriminate the two regions. From this analysis, 20 *m/z* values had AUC values of >0.5 which were then investigated further if they have significant abundances in the CRLM tissues (Table 2). The 20 *m/z* values were observed between a *m/z* range of 1000 to 2300 (Fig. 3A-B). Subsequently, a ROC plot with associated AUC values for each peptide was generated to plot the specificity and sensitivity in relation to tumour and stroma. Of the 20 *m/z* values, *m/z* 1589.876 with 0.618 AUC (95% CI: 0.563-0.674), and *m/z* 1092.727 with 0.562 AUC (95% CI: 0.504-0.620) were found to be high in intensity in tumour regions relative to stroma regions. Unfortunately, *m/z* 1589.876 and *m/z* 1092.727 could not be identified from the LC-MS/MS data (Fig. 3C-F). The remaining 18 *m/z* values with AUC values >0.5 were not statistically significant (Table 2 and Supp. Fig 1).

**Figure 3.**
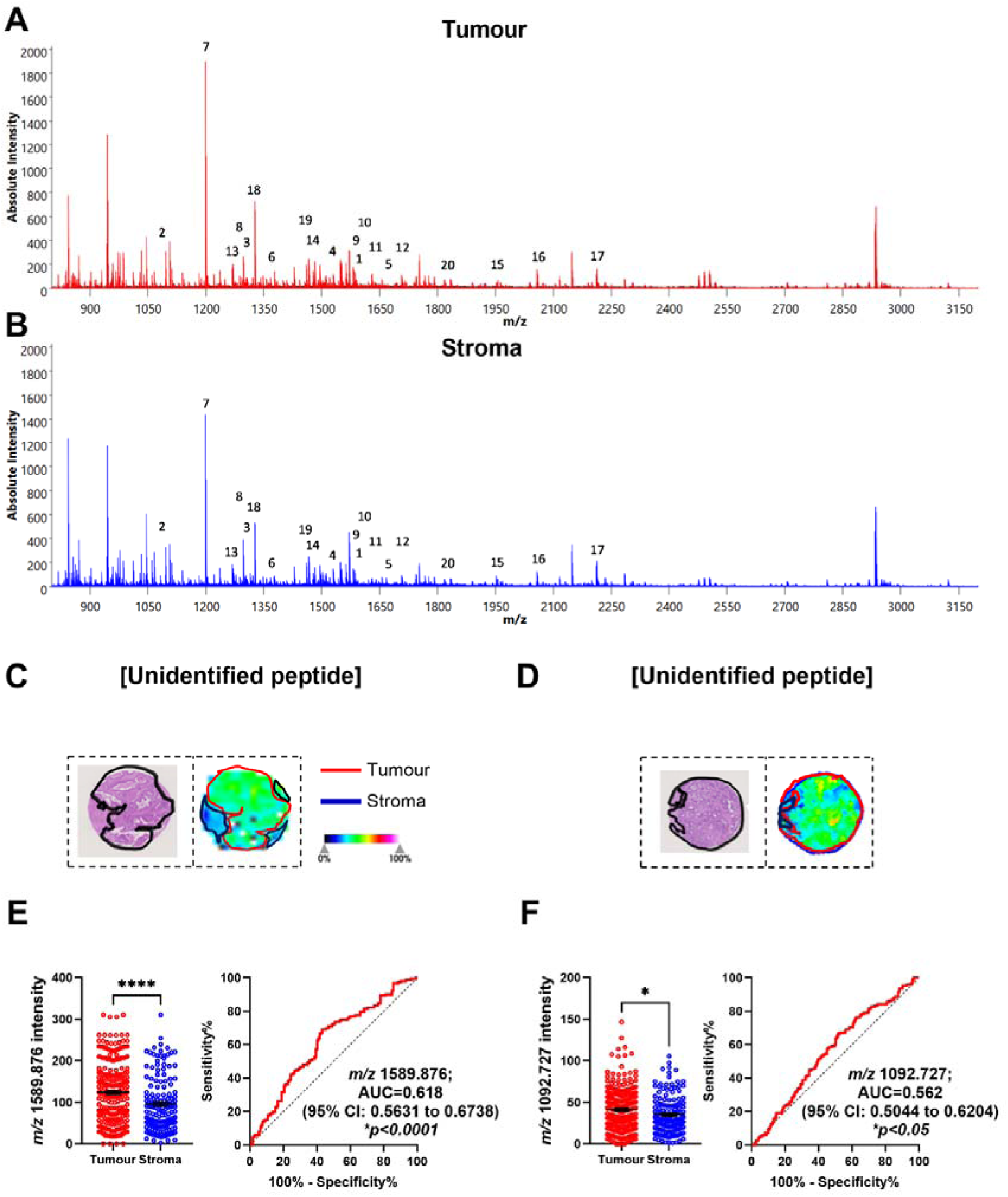
Identification of tryptic peptides associated with CRLM tumour regions using MALDI-MSI and LC-MS/MS. (A) Sum spectra of CRLM tumour (red) and (B) stroma (blue) in CRLM tumour cores. (C, D) H&E images (left) of a representative tumour core with tumour regions annotated in black and ion intensity maps (right) of most discriminative *m/z* values between tumour (red) and stroma (red). Tumour cores are 1.5mm in diameter. Bar graphs showing mean ± SEM and ROC plots for (E) *m/z* 1589.876 and (F) *m/z* 1092.727. *\*p<0.05, ****p<0.0001.* Student’s t test. Each dot represents a single tumour core.

**Table 2.**
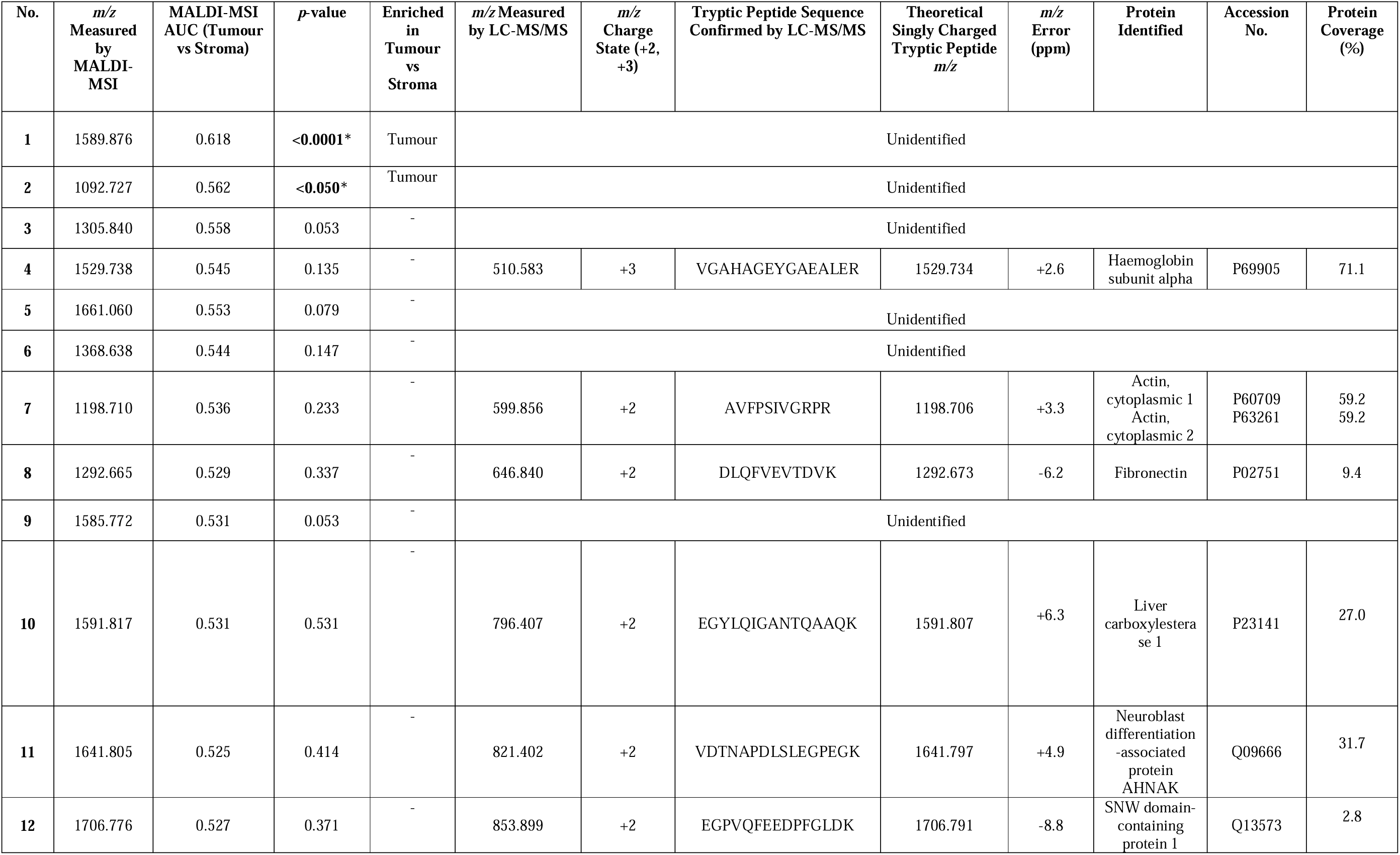
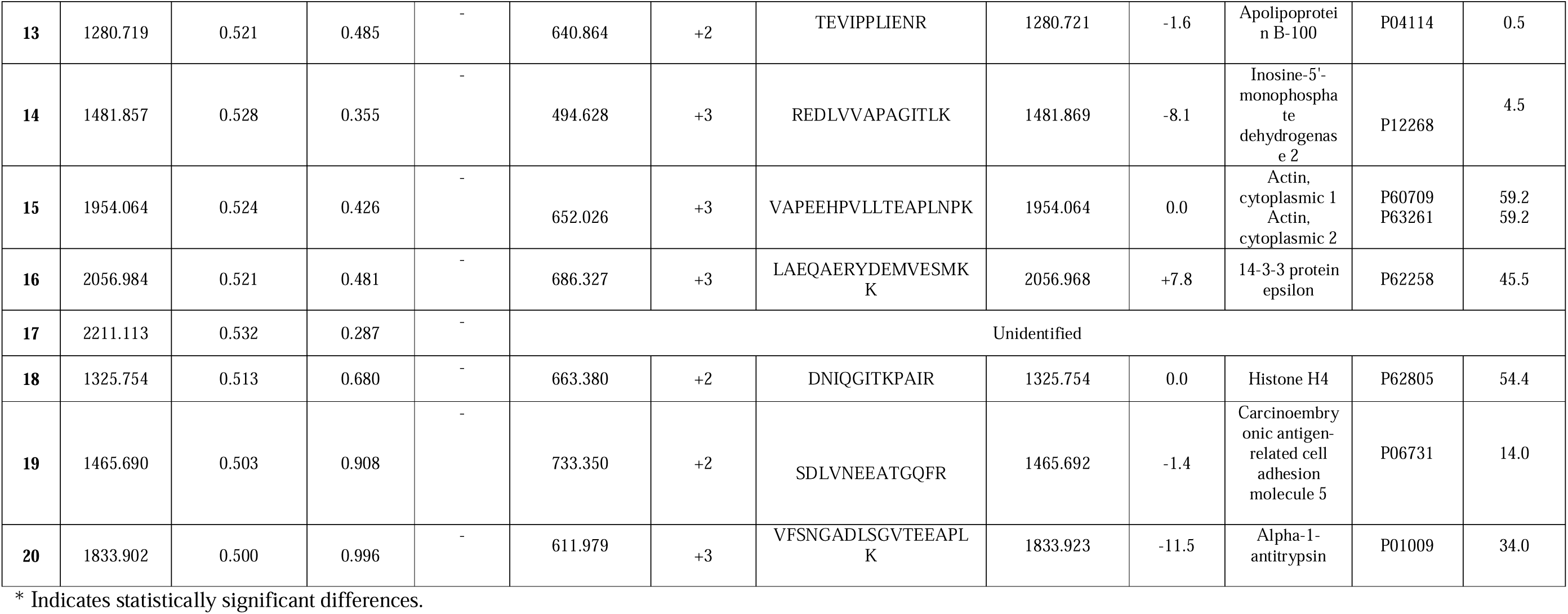
Tryptic peptides and proteins that discriminate between tumour and stroma regions in CRLM by MALDI-MSI and LC-MS/MS.

### Identification of tryptic peptides associated with biological sex

Next, we determined if there were differences in the spatial proteome between male and female CRLM tumour tissues. Selecting, MALDI-MSI peaks with an AUC of >0.5 resulted in the identification of 14 *m/z* values. From this analysis, 8 *m/z* values (*m/z* 2211.113, 1296.682, 1570.676, 1669.829, 1461.702, 1465.697, 1235.618, and 1366.628) were found to be significantly different in the tumour region between males and females with AUC values between 0.600-0.695 (Table 3 and Supp. Fig. 2). Interestingly, we identified 4 (*m/z* 2211.113, 1296.682, 1570.676, 1366.628) to be higher in intensity in females (Fig 4A-D), whereas another 4 (*m/z* 1669.829, 1461.702, 1465.697, 1235.618) were higher in intensity in males (Fig. 4E-H). In the stroma (Supp. Fig. 3), 8 (*m/z* 2211.113, 1296.682, 1570.676, 1669.829, 1465.697, 1366.628, 1661.060 and 1305.840) were identified to be statistically different between male and females, with 6 (*m/z* 2211.113, 1296.682, 1570.676, 1366.628, 1661.060 and 1305.840) higher in females (Fig 5A-F) compared to 2 (*m/z* 1669.829 and 1465.697) in males (Fig 5G-H). The AUC values ranged from 0.633-0.677.

**Figure 4.**
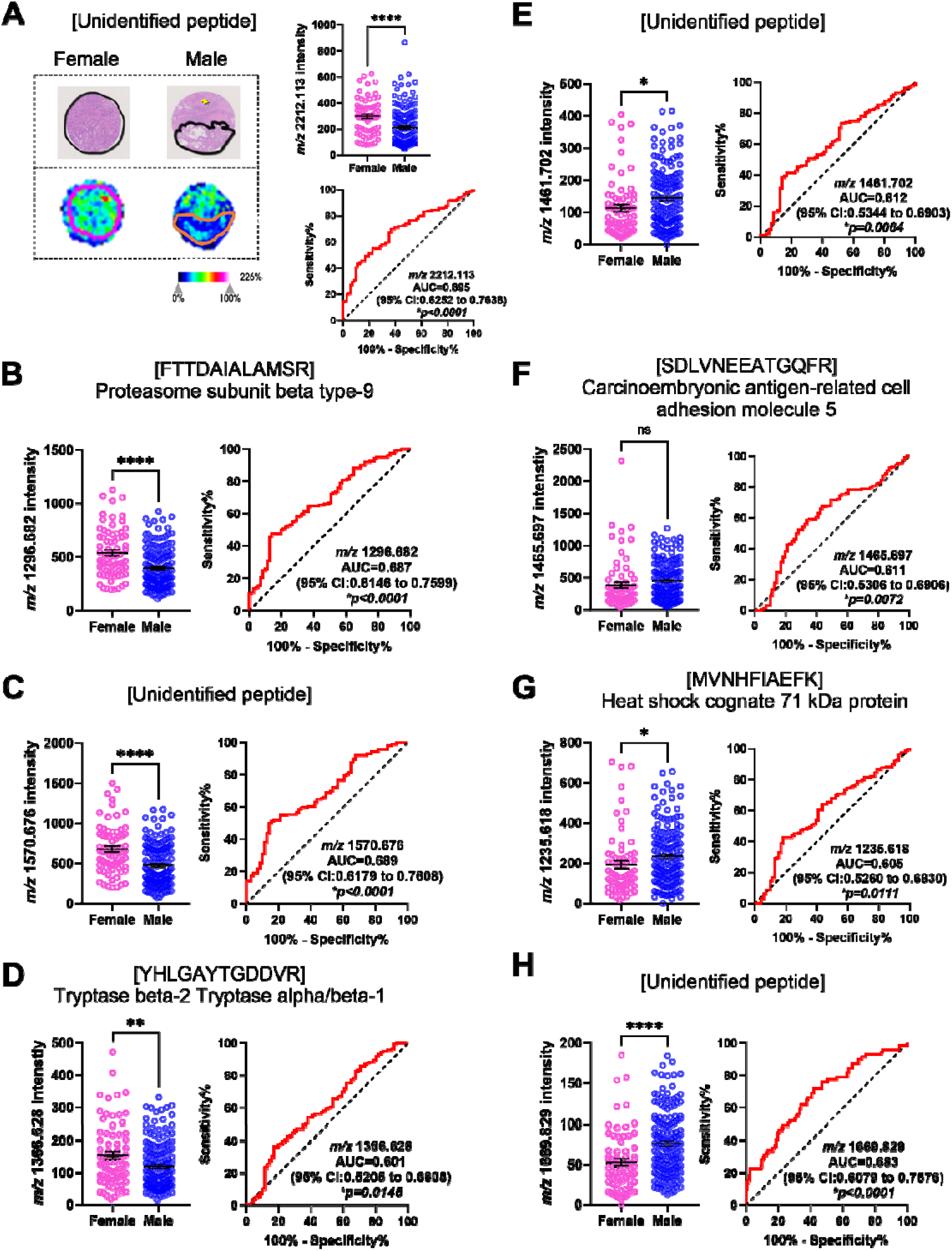
Significantly abundant tryptic peptides associated with male and female CRLM tumour regions, by MALDI-MSI and LC-MS/MS. (A) Representative H&E and ion intensity map of tumour region annotated CRLM tumour cores for *m/z* 2211.113. Tumour cores are 1.5mm in diameter. (A-D) Tryptic peptides (*m/z* 2211.113, 1296.682, 1570.676, and 1366.628) significantly more abundant in females relative to males. Tryptic peptides were identified as (A) unidentified, (B) Proteasome subunit beta type-9, (C) unidentified, and (D) Tryptase beta-2 Tryptase alpha/beta-1. (E-H) Tryptic peptides (*m/z* 1669.829, 1461.702, 1466.697, and 1235.618) significantly more abundant in males relative to females. These tryptic peptides were identified as (E) unidentified, (F) Carcinoembryonic antigen-related cell adhesion molecule 5, (G) Heat shock cognate 71 kDa protein, and (H) unidentified. Bar graphs showing mean ± SEM and ROC plots are presented. *\*\*\*\*p<0.0001.* Student’s t test. Each dot represents a single tumour core.

**Figure 5.**
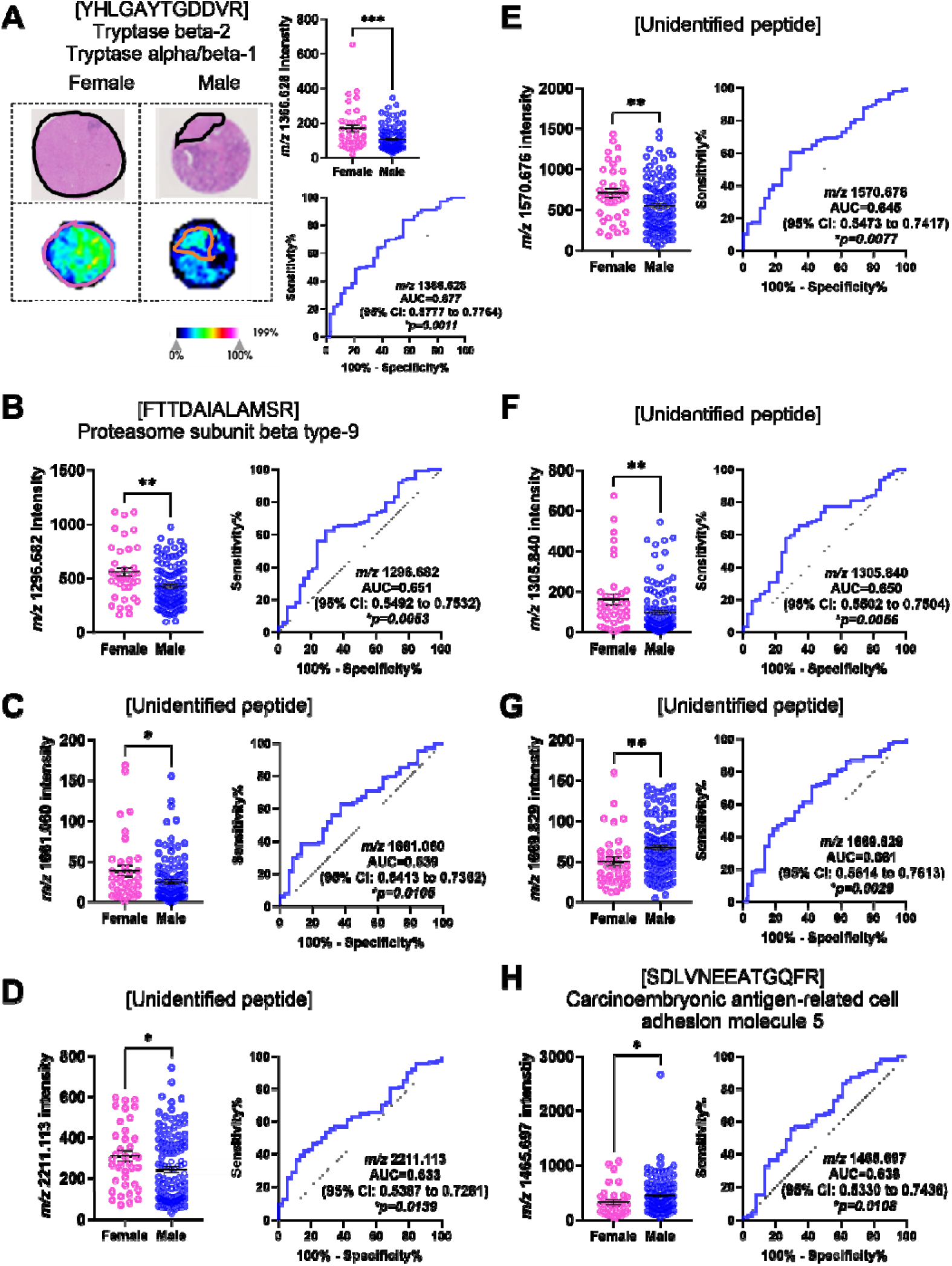
Significantly abundant tryptic peptides associated with male and female CRLM stroma regions, by MALDI-MSI and LC-MS/MS. (A) Representative H&E and ion intensity map of stroma region annotated CRLM tumour cores for *m/z* 1366.628. Tumour cores are 1.5mm in diameter. (A-F) Tryptic peptides (*m/z* 1366.628, 1296.682, 2211.113, 1570.676, 1661.060 and 1305.840) significantly more abundant in females relative to males. Tryptic peptides were identified as (A) Tryptase beta-2 Tryptase alpha/beta-1, (B) Proteasome subunit beta type-9, and (C-F) unidentified. (G-H) Tryptic peptides (*m/z* 1669.829 and 1461.702) significantly more abundant in males relative to females. These tryptic peptides were identified as (G) unidentified and (H) Carcinoembryonic antigen-related cell adhesion molecule 5. Bar graphs showing mean ± SEM and ROC plots are presented. *\*\*\*\*p<0.0001.* Student’s t test. Each dot represents a single tumour core.

**Table 3.** Tryptic peptides and proteins that discriminate between biological sex in CRLM by MALDI-MSI and LC-MS/MS.

| No. | *m/z*<br>Measure<br>d by<br>MALDI-<br>MSI | MALDI-<br>MSI<br>AUC<br>Stroma<br>(Male vs<br>Female) | *p*-value | Stroma<br>Enriched<br>(Male vs<br>Female) | MALDI-<br>MSI<br>AUC<br>Tumour<br>(Male vs<br>Female) | *p*-value | Tumour<br>Enriched<br>(Males vs<br>Female) | *m/z*<br>Measure<br>d by LC-MS/MS | *m/z*<br>Char<br>ge<br>State<br>(+2,<br>+3) | Tryptic Peptide<br>Sequence<br>Confirmed by LC-MS/MS | Theoretic<br>al Singly<br>Charged<br>Tryptic<br>Peptide<br>*m/z* | *m/z*<br>Error<br>(ppm) | Protein<br>Identified | Accessio<br>n No. | Protein<br>Coverag<br>e (%) |
| --- | --- | --- | --- | --- | --- | --- | --- | --- | --- | --- | --- | --- | --- | --- | --- |
| 1 | 2211.113 | 0.633 | **0.0139\*** | Female | 0.695 | **<0.0001\*** | Female | Unidentified |  |  |  |  |  |  |  |
| 2 | 1296.682 | 0.662 | **0.0053\*** | Female | 0.687 | **<0.0001\*** | Female | 648.834 | +2 | FTTDAIALAMSR | 1296.662 | +15.4 | Proteasome<br>subunit beta<br>type-9 | P28065 | 18.30 |
| 3 | 1570.676 | 0.645 | **0.0077\*** | Female | 0.689 | **<0.0001\*** | Female | Unidentified |  |  |  |  |  |  |  |
| 4 | 1669.829 | 0.661 | **0.0029\*** | Male | 0.683 | **<0.0001\*** | Male | Unidentified |  |  |  |  |  |  |  |
| 5 | 1461.702 | 0.563 | 0.2489 | - | 0.612 | **0.0064\*** | Male | Unidentified |  |  |  |  |  |  |  |
| 6 | 1465.697 | 0.638 | **0.0108\*** | Male | 0.611 | **0.0072\*** | Male | 733.350 | +2 | SDLVNEEATGQF<br>R | 1465.692 | +3.4 | Carcinoembry<br>onic antigen-related cell<br>adhesion<br>molecule 5 | P06731 | 14.0 |
| 7 | 1235.618 | 0.582 | 0.132 | - | 0.605 | **0.0111\*** | Male | 618.316 | +2 | MVNHFIAEFK | 1235.624 | -4.9 | Heat shock<br>cognate 71<br>kDa protein | P11142 | 31.30 |
| 8 | 1366.628 | 0.677 | **0.0011\*** | Female | 0.601 | **0.0145\*** | Female | 683.823 | +2 | YHLGAYTGDDV<br>R | 1366.639 | -8.0 | Tryptase beta-2<br>Tryptase<br>alpha/beta-1 | P20231<br>Q15661 | 24<br>24 |
| 9 | 1481.857 | 0.558 | 0.2874 | - | 0.551 | 0.2128 | - | 494.628 | +3 | REDLVVAPAGIT<br>LK | 1481.869 | -8.1 | Inosine-5'-<br>monophosphat<br>e<br>dehydrogenase<br>2 | P12268 | 4.5 |
| 10 | 1280.719 | 0.549 | 0.3971 | - | 0.550 | 0.2247 | - | 640.864 | +2 | TEVIPPLIENR | 1280.721 | -1.6 | Apolipoprotein<br>B-100 | P04114 | 0.5 |
| 11 | 1441.676 | 0.557 | 0.2972 | - | 0.547 | 0.2542 | - | 721.343 | +2 | EFTPQMQAAYQ<br>K | 1441.678 | -1.4 | Hemoglobin<br>subunit delta | P02042 | 40.10 |
| 12 | 1661.060 | 0.639 | **0.0105\*** | Female | 0.520 | 0.6212 | - | Unidentified |  |  |  |  |  |  |  |
| 13 | 1305.840 | 0.650 | **0.0056\*** | Female | 0.505 | 0.8958 | - | Unidentified |  |  |  |  |  |  |  |
| 14 | 1529.738 | 0.557 | 0.2972 | - | 0.502 | 0.9697 | - | 510.583 | +3 | VGAHAGEYGAE<br>ALER | 1529.734 | +2.6 | Hemoglobin<br>subunit alpha | P69905 | 71.1 |

\* Indicates statistically significant differences.

In total 10 unique *m/z* values were identified to discriminate between male and female tissues (Table 3). Six of the 10 *m/z* values were associated with biological sex regardless of the region type (tumour or stroma). Using LC-MS/MS, we identified 3 peptides, [FTTDAIALAMSR] (*m/z* 1296.682) from Proteasome subunit beta type-9 (PSMB9), [SDLVNEEATGQFR] (*m/z* 1465.697) from Carcinoembryonic antigen-related cell adhesion molecule 5 (CEACAM5), and [YHLGAYTGDDVR] (*m/z* 1366.628) from Tryptase beta-2 (TPSB2) or Tryptase alpha/beta-1 (TPSAB1), while the corresponding peptides for *m/z* 2211.113, *m/z* 1570.676 and *m/z* 1669.829 could not be identified. Further, an unidentified *m/z* value (*m/z* 1461.702) and [MVNHFIAEFK] (*m/z* 1235.618) from Heat shock cognate 71 kDa protein (HSPA8) were higher in male tumour regions, while two unidentified *m/z* values (1661.060 and 1305.840) were higher in female stroma regions. Together, these results indicated that biological sex could contribute to changes to both the overall and region-specific tumour proteome.

### Identification tryptic peptides linked to survival

To identify potential prognostic markers for CRLM, we classified our patient cohort into 3 groups based on survival (0-3 years, 4-6 years, and 7+ years). Specific to the tumour region (Supp. Fig 4), we identified 6 *m/z* values that had an AUC >0.5 between 0-3 and 7+ years survival. From this list, 5 *m/z* values (*m/z* 1325.754, 1529.738, 1481.857, 1305.840 and 1661.060) achieved statistical significance (Table 4). [DNIQGITKPAIR] (*m/z* 1325.754) from histone H4 had the highest AUC value of 0.720 (95% CI: 0.6320-0.8086) (Fig. 6A). Additionally, [VGAHAGEYGAEALER] (*m/z* 1529.738) from Haemoglobin subunit alpha (HBA1), [REDLVVAPAGITLK] (*m/z* 1481.857) from Inosine-5’-monophosphate dehydrogenase 2 (IMPDH2), and 2 unidentified *m/z* values (*m/z* 1305.840 and 1661.060) were all relatively high abundant in patients who had poor survival outcomes (0-3 years post-surgery) compared to those who have survived for 7 or more years (Fig. 6). Analysing the stromal regions (Supp. Fig. 5), only histone H4 replicated the tumour analysis showing higher abundance in patients with poor (0-3 years) survival outcomes (Supp. Fig. 6). Together, our MALDI-MSI analysis identified peptides that associated with duration of survival in CRLM patients.

**Figure 6.**
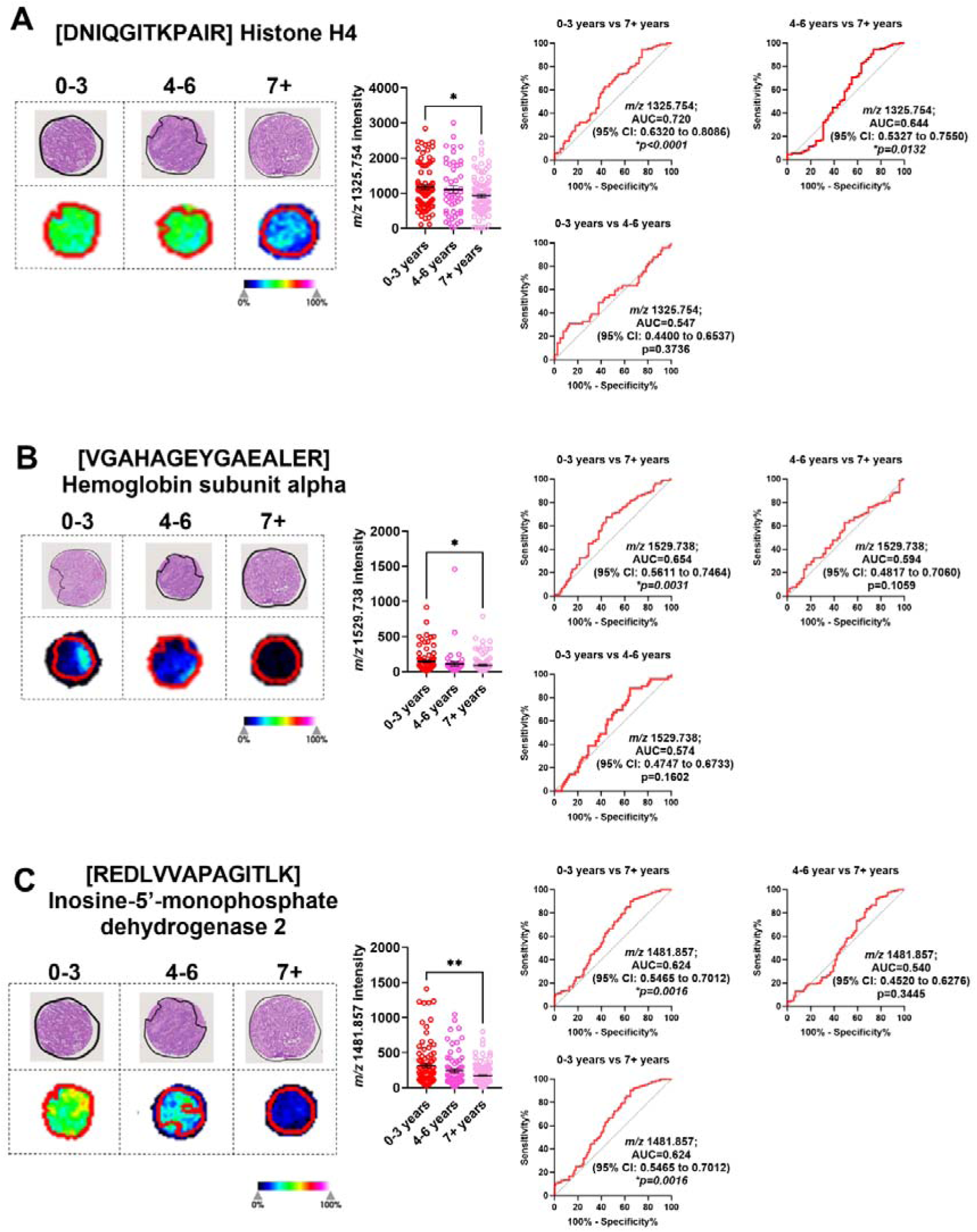
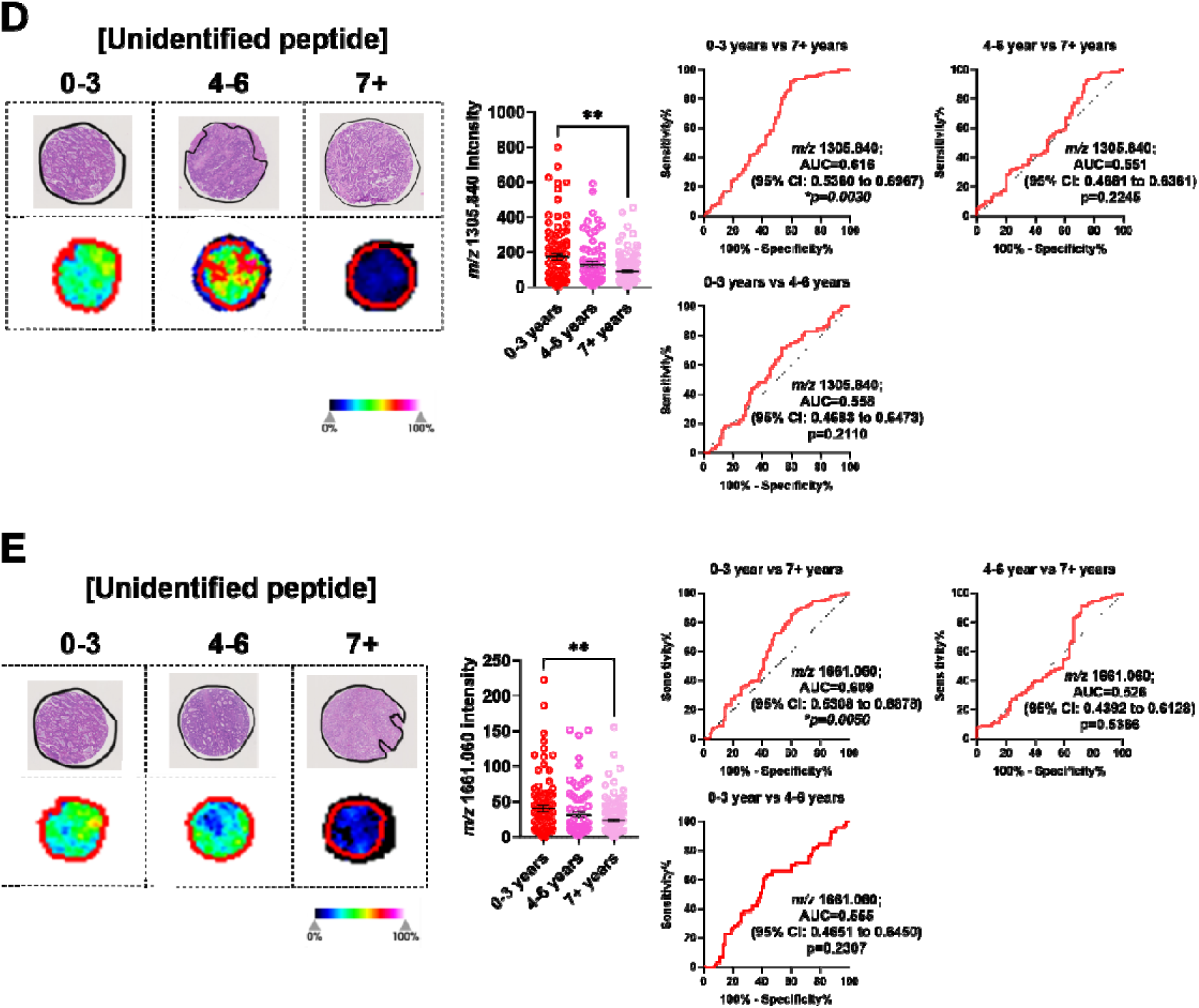
Significantly abundant tryptic peptides associated with poor overall survival (0-3 years) in CRLM tumour regions. CRLM cores were subdivided based on overall survival after curative-intent surgery (0-3, 4-6, and 7+ years). Representative H&E and ion intensity maps of tumour region annotated CRLM tumour cores for *m/z* 1325.754, 1529.738, 1481.857, 1305.840, and 1661.060. Tumour cores are 1.5mm in diameter. These tryptic peptides were identified as (A) Histone H4, (B) Haemoglobin subunit alpha, (C) Inosine-5’-monophosphase dehydrogenase 2 and (D-E) unidentified by LC-MS/MS. Bar graphs showing mean ± SEM and ROC plots are presented. *\*p<0.05*, *\*\*p<0.01.* Ordinary one-way ANOVA. Each dot represents a single tumour core.

**Table 4.** Tryptic peptides and proteins that discriminate overall survival identified by MALDI-MSI and LC-MS/MS.

| No. | *m/z*<br>Measured<br>by<br>MALDI-<br>MSI | MALDI-<br>MSI<br>AUC<br>Tumour<br>(0-3 vs<br>7+<br>Years) | *p*-value | MALDI-<br>MSI<br>AUC<br>Tumour<br>(0-3 vs<br>4-6<br>Years) | *p*-<br>value | MALDI-<br>MSI<br>AUC<br>Tumour<br>(4-6 vs<br>7+<br>Years) | *p*-value | *m/z*<br>Measured<br>by LC-MS/MS | *m/z*<br>Charge<br>State<br>(+2,<br>+3) | Tryptic Peptide<br>Sequence Confirmed by<br>LC-MS/MS | Theoretical<br>Tryptic<br>Peptide<br>*m/z* | *m/z*<br>Error<br>(ppm) | Protein<br>Identified | Accession<br>No. | Protein<br>Coverage<br>(%) |
| --- | --- | --- | --- | --- | --- | --- | --- | --- | --- | --- | --- | --- | --- | --- | --- |
| 1 | 1325.754 | 0.720 | <0.0001* | 0.547 | 0.3736 | 0.644 | 0.0132* | 442.589 | +3 | DNIQGITKPAIR | 1325.754 | 0.0 | Histone H4 | P62805 | 54.4 |
| 2 | 1529.738 | 0.654 | 0.0031* | 0.574 | 0.1602 | 0.594 | 0.1059 | 765.371 | +2 | VGAHAGEYGAEALER | 1529.734 | +2.6 | Hemoglobin<br>subunit alpha | P69905 | 71.1 |
| 3 | 1481.857 | 0.624 | 0.0016* | 0.573 | 0.1157 | 0.540 | 0.3445 | 494.628 | +3 | REDLVVAPAGITLK | 1481.869 | -8.1 | Inosine-5'-<br>monophosphate<br>dehydrogenase<br>2 | P12268 | 4.5 |
| 4 | 1305.840 | 0.616 | 0.003* | 0.558 | 0.211 | 0.551 | 0.2245 | Unidentified |  |  |  |  |  |  |  |
| 5 | 1661.060 | 0.609 | 0.005* | 0.555 | 0.2307 | 0.526 | 0.5366 | Unidentified |  |  |  |  |  |  |  |
| 6 | 1589.876 | 0.594 | 0.0696 | 0.552 | 0.3281 | 0.665 | 0.0045* | Unidentified |  |  |  |  |  |  |  |

\* Indicates statistically significant differences.

## Discussion

CRLM is a highly lethal and heterogeneous disease with poor prognostic tests available [26, 27]. While previous studies have reported on the use of MALDI-MSI in CRLM tissue, most have used small sample sizes to only describe the proteomic landscape of CRLM [19, 20]. To the best of our knowledge, this study is the first study to utilise MALDI-MSI combined with LC-MS/MS to identify potential protein biomarkers in CRLM patients. Furthermore, we applied this technology on the largest cohort of CRLM patients collected in Australia.

The tumour-stroma ratio is a well-known independent prognostic indicator for CRC, with high levels of stroma associating with poor survival [28, 29]. Thus, we determined if there are consensus proteins that could discriminate between the two regions in our heterogenous CRLM sample cohort by MALDI-MSI. MALDI-MSI analysis between tumour and intratumoral stroma regions, identified peptides that associate with tumour regions. Among the 20 *m/z* values identified using MALDI-MSI, 2 *(m/z* 1589.876 and 1092.727) reached statistical significance, however it was not possible to establish matches with our LC-MS/MS data. Nevertheless, our MALDI-MSI data was able to find features that distinguish tumours from stroma.

While it is widely appreciated that biological sex can greatly affect cancer biology and treatment outcomes [30, 31], proteomic analyses that describe such contributions, especially in CRLM, are rarely reported. In this study, we investigated if biological sex was associated with differences in the spatial proteome of CRLM. We identified PSMB9, CEACAM5 and TPSB2/TPSAB1 and 3 unidentified *m/z* values (*m/z* 2211.113, 1570.676 and 1669.829) that had significant differences in abundance between male and female CRLM tissues. These were sex-specific for both tumour and stroma regions. Interestingly, HSPA8 and an unidentified *m/z* value (*m/z* 1461.702) localised to tumours specific to males, and 2 unidentified *m/z* values (*m/z* 1305.840 and 1661.060) localised to stroma, that were specific to females. Together these results show for the first time that biological sex may have an underappreciated role in the spatial proteome of CRLM that may have currently unknown consequences to its pathophysiology. Further research is required to determine if expression of these proteins affect sex specific CRLM progression and treatment outcomes.

We report that histone H4, HBA1, IMPDH2, and two unidentified *m/z* values (*m/z* 1305.840 and 1661.060) are more abundant in tumour tissues of CRLM patients with poor survival. In this study, only histone H4 expression in tumour and/or stroma was enriched in patients that survived for >3 years. Histones are key epigenetic regulators. Their post-translational modifications (acetylation, methylation and phosphorylation) can be linked with gene expression changes that lead to cancer progression including CRC and hence are being investigated for their therapeutic and prognostic potential [32]. Histone H4 upregulation has been linked to platinum-based chemotherapy resistance in malignancies [33] and may explain its association with poor prognosis in our patient cohort. Interestingly, histone H4 was identified to be elevated in CRC patient plasma by mass spectrometry (detecting the [DNIQGITKPAI] peptide sequence) [34], and ELISA [35]. Thus, histone H4 could potentially be developed as a blood biomarker to identify CRLM patients with worse prognostic outcomes. The relationship between histone H4 tumour tissue expression, plasma levels, and survival outcomes is unknown and warrants further investigation. HBA1 is the alpha subunit of hemoglobulin. Anaemia, measured by blood hemoglobulin levels, has been linked with tumour hypoxia and poor outcomes in solid tumours [36]. However, the specific contribution of HBA1 in cancer progression is unknown. Recent single cell RNA-sequencing analysis of gastric cancers shows that HBA1 is overexpressed in gastric cancer cells. Importantly, patients with high tumour HBA1 expression led to poor overall survival supporting our findings in CRLM [37]. The contributions of IMPDH2 in CRC progression is well established. IMPDH2 is an isoform of IMPDH, an enzyme critical for biosynthesis of purine nucleotides and is essential for DNA synthesis [38]. IMPDH2 is commonly upregulated in malignancies [39] including primary CRC [40]. IMPDH2 has been shown to promote CRC tumorigenesis [41], metastatic potential [42] and in methotrexate resistance [43]. Thus, IMPDH2 has been previously implied as a potential prognostic marker for CRC outcomes and may be useful as a drug target. Collectively, our study identified previously reported and novel tryptic peptides candidates that may be able to predict poor outcomes in CRLM patients. Considering the reported function of these proteins in cancer, these may also be used as novel targets in CRLM therapy development.

In this study we used a manual indirect identification approach, where we matched the MALDI-MSI data with data from an independent LC-MS/MS experiment. As part of this workflow, we have implemented internal calibrants to allow high accuracy peptide matching [24]. However, there are limitations in this study to consider, including the identification of peptides with the similar *m/z* values which fit within the same MALDI-MSI mass tolerance window, therefore potentially resulting in mis-assignments. Additional validation by *in situ* tandem MS fragmentation to confirm the identity of the peptides may improve peptide identification [44]. However, this approach is only feasible for peptides with high signal intensity. Recently, several automated annotation tools have been developed, which even include post-translational modifications [24, 45]. These tools might be used to identify more of the 471 peptides from the LC-MS/MS data. Moreover, additional methods such as immunohistochemistry could be employed to validate the abundance of the identified proteins in the metastatic tumour regions. A second independent cohort of patients is required to validate our findings and confirm the prognostic potential of the identified proteins that associated with poor CRLM survival outcomes. Nonetheless, this study provides novel and valuable insights into CRLM biology and identifies potential prognostic biomarkers and targets that are needed in CRLM.

## Conclusions

In summary, this is the first study to utilise MALDI-MSI with LC-MS/MS to identify potential spatial proteomic tissue biomarkers in the largest cohort of Australian CRLM patients to date. Using these workflows, spatial proteomic features present in CRLM were revealed and changes within the tumour proteome identified in relation to clinical features, including sex and overall survival. Importantly, we reveal that biological sex could impact the spatial proteome profile of CRLM tumours and identified several proteins that associate with poor survival in CRLM patients. These findings should be further explored to identify new prognostic markers and therapeutic targets that are highly needed in CRLM.

## Statements and Declarations

### Funding

This work was supported by a Cancer Council SA Beat Cancer Infrastructure grant with matched funding support from The Hospital Research Foundation Group (GM. and K.F.). K.F. was supported by a The Hospital Research Foundation Group Early Career Fellowship. C.L. was supported by a University of Adelaide Postgraduate Research Scholarship. The authors acknowledge Bioplatforms Australia, the University of South Australia, and the State and Federal Governments, which co-fund the NCRIS-enabled Mass Spectrometry and Proteomics facility at the University of South Australia.

### Competing Interests

The authors have no relevant financial or non-financial interests to disclose.

### Author Contributions

Conceptualisation C.L., M.B., M.K.H and K.F.; Collection and/or assembly of data: C.L., M.B., Y.L., T.T., C.Y., J.P., and G.K. Data Analysis and Interpretation: C.L., M.B., Y.L., C.Y., M.K.H, and K.F., resources: J.P., G.M., P.H, and K.F.; writing—original draft preparation, C.L., M.B., P.D., M.K.H and K.F.; writing—review and editing, C.L., M.B., C.Y., P.D., M.K.H. and K.F.; supervision, P.H., G.M., M.K.H, and K.F., funding acquisition, C.L., G.M., M.K.H and K.F.; All authors have read and agreed to the published version of the manuscript.

### Data Availability

The data underlying this article are available in this published article and its supplementary information files. Additional data are available upon request to the corresponding author.

### Ethics Approval

This study was approved by the Human Research Ethics Committee of the Central Adelaide Local Health Network under protocol number 12237.

### Consent to Participate

Samples collected were retrospective patient tissues and was not consented as advised by our Human Research Ethics Committee.

## Notes

### Competing Interest Statement

The authors have declared no competing interest.

