## Supplementary File for "Use of tryptic peptide MALDI mass spectrometry imaging to identify the spatial proteomic landscape of colorectal cancer liver metastases"

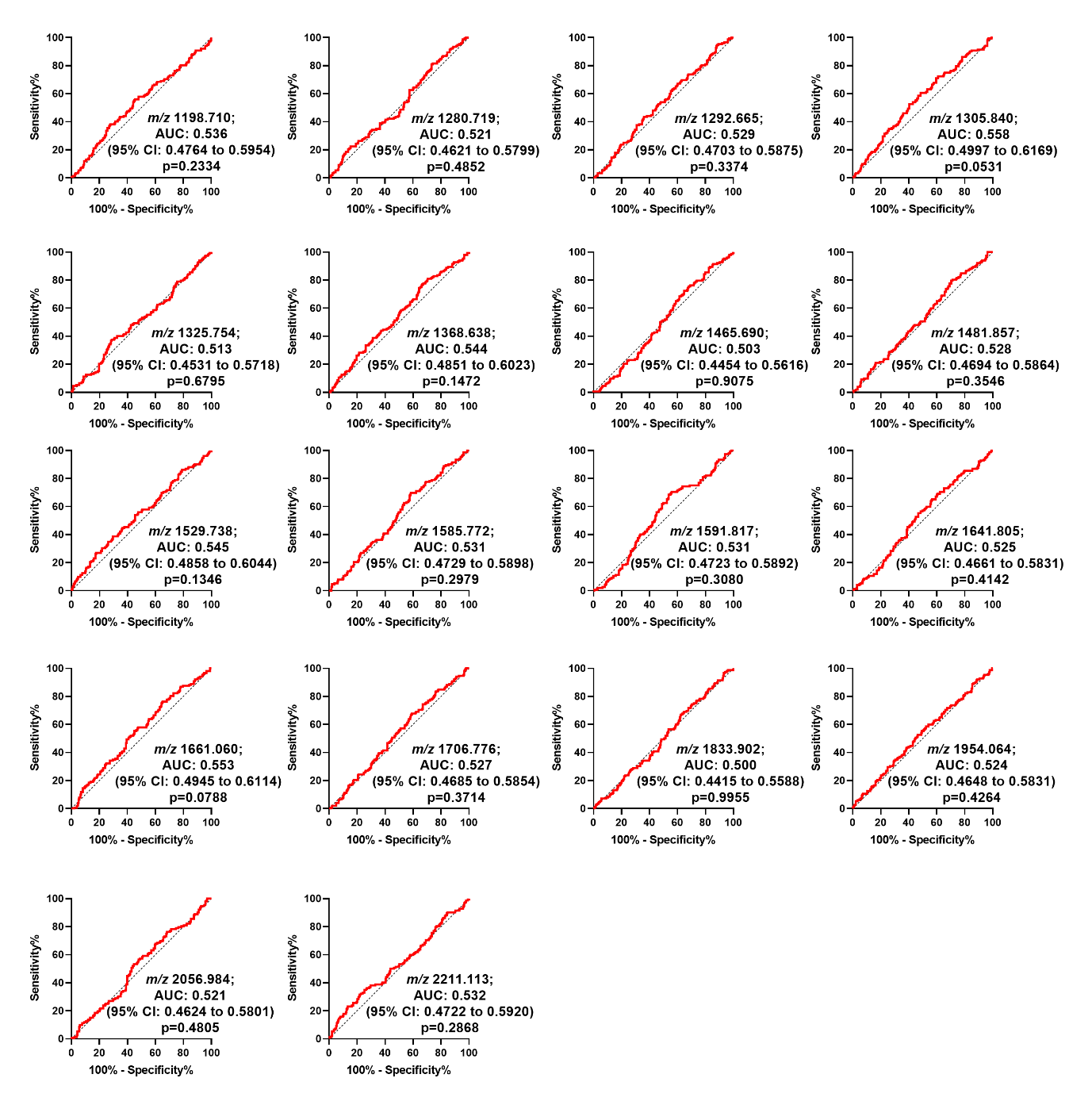

**Supplementary Figure 1.** ROC plots with AUC values >0.5 for tryptic peptides (*m/z*) that discriminate between tumour and stroma within CRLM tumour cores. Corresponds to Table 2.

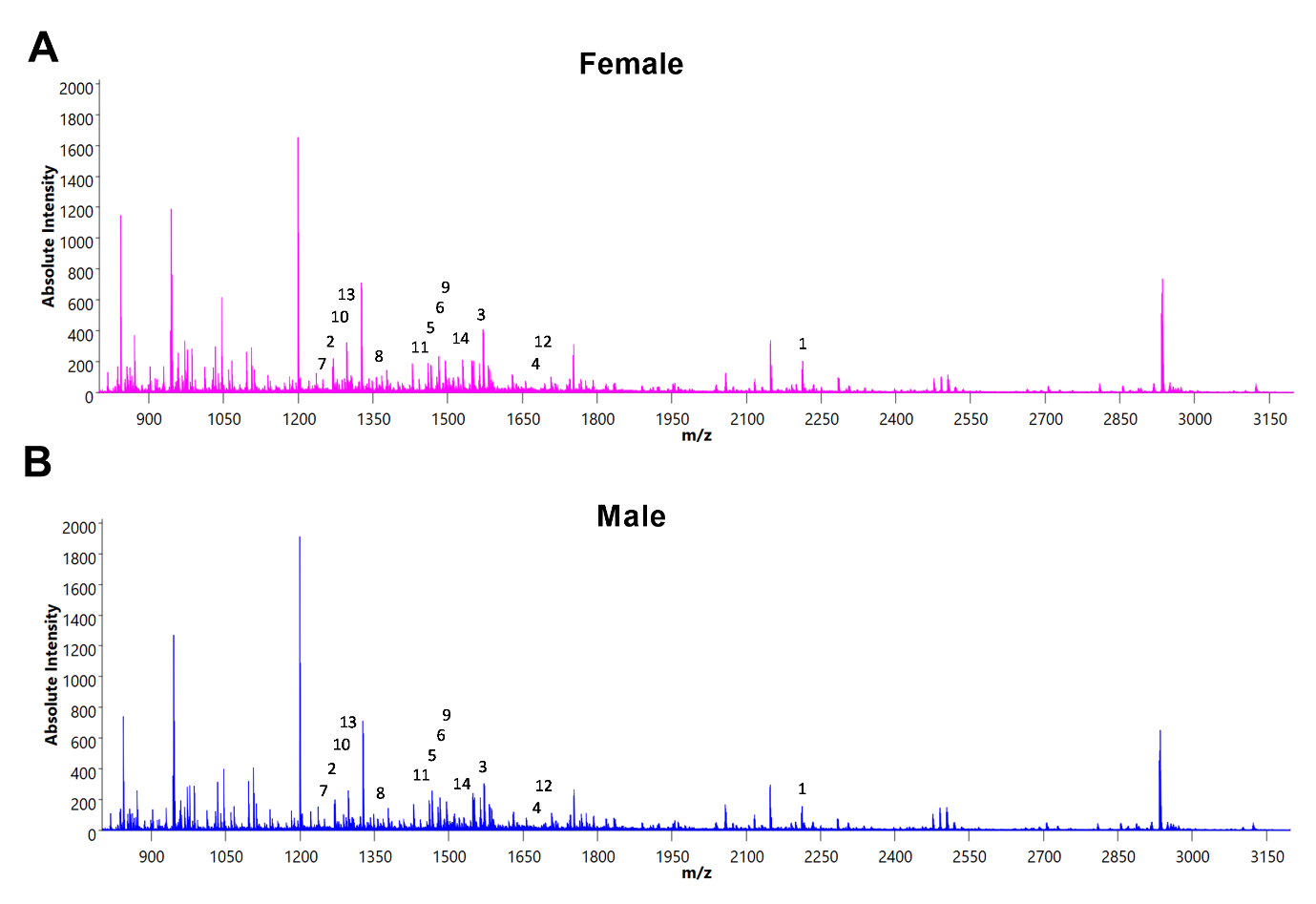

**Supplementary Figure 2.** Overall spectrum based on biological sex in tumour. (A) Female, and (B) Male.

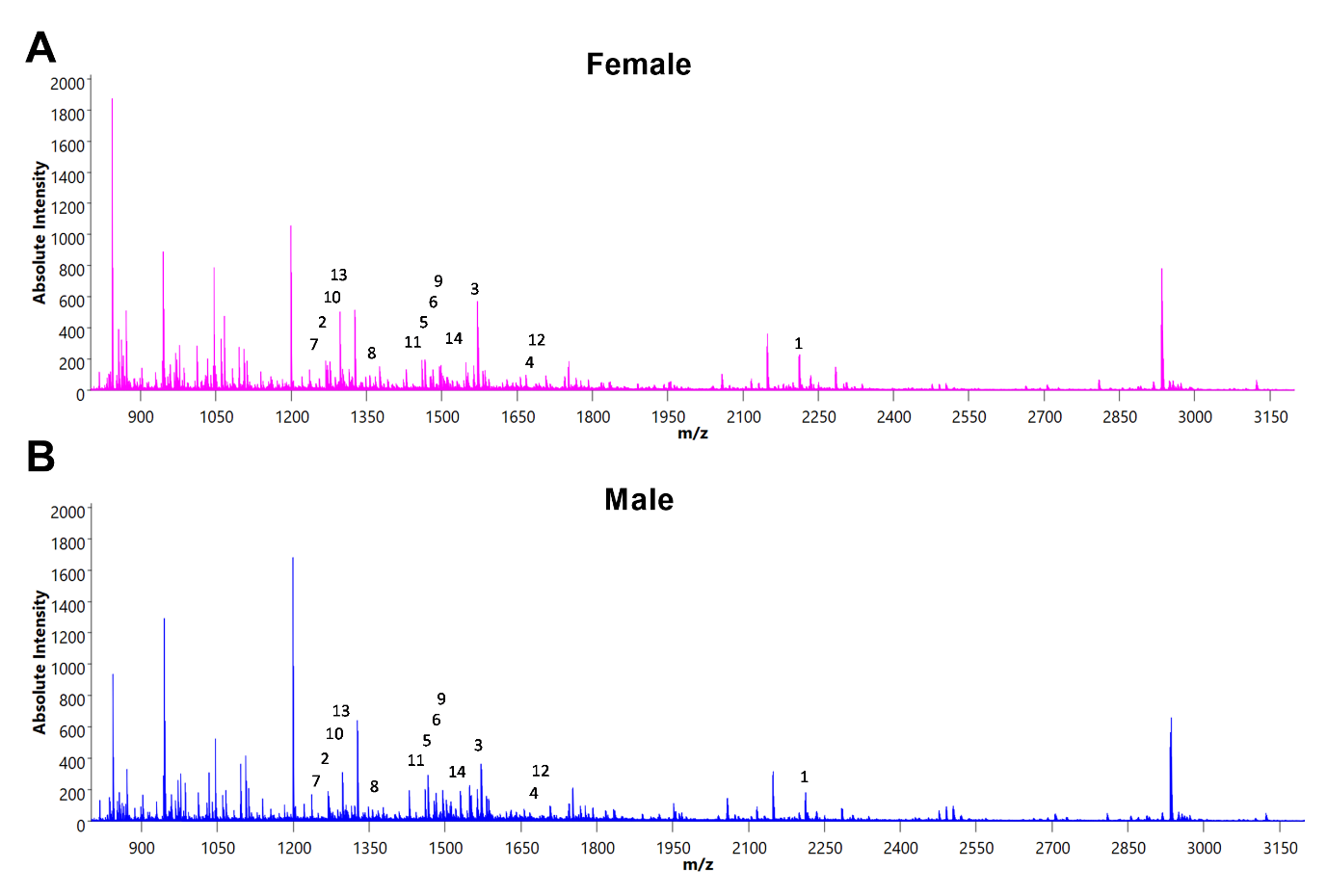

**Supplementary Figure 3.** Overall spectrum based on biological sex in stroma. (A) Female, and (B) Male.

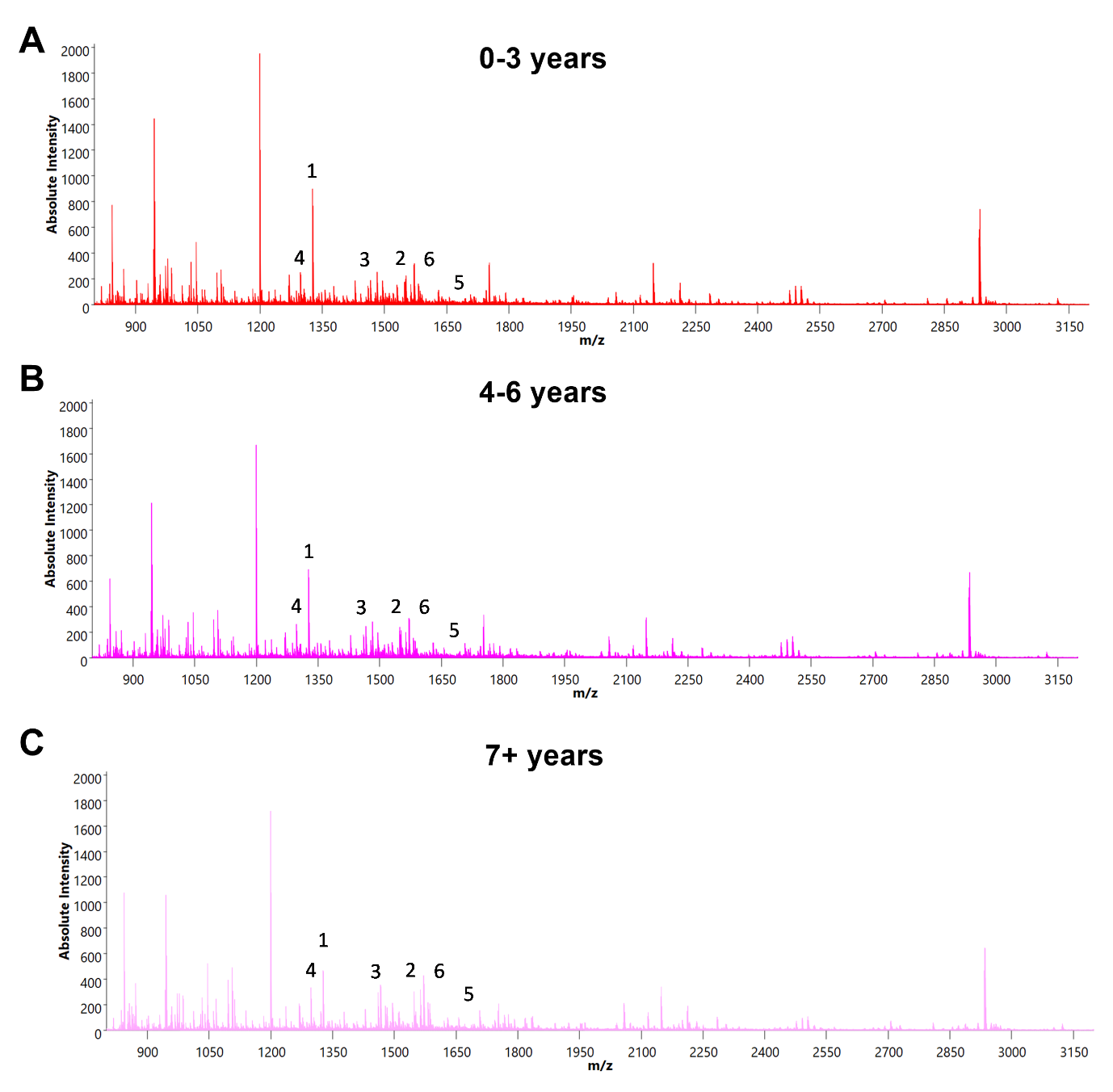

**Supplementary Figure 4.** Overall spectrum based on the three overall survival groups in tumour. (A) 0-3 years, (B) 4-6 years, and (C) 7+ years survival.

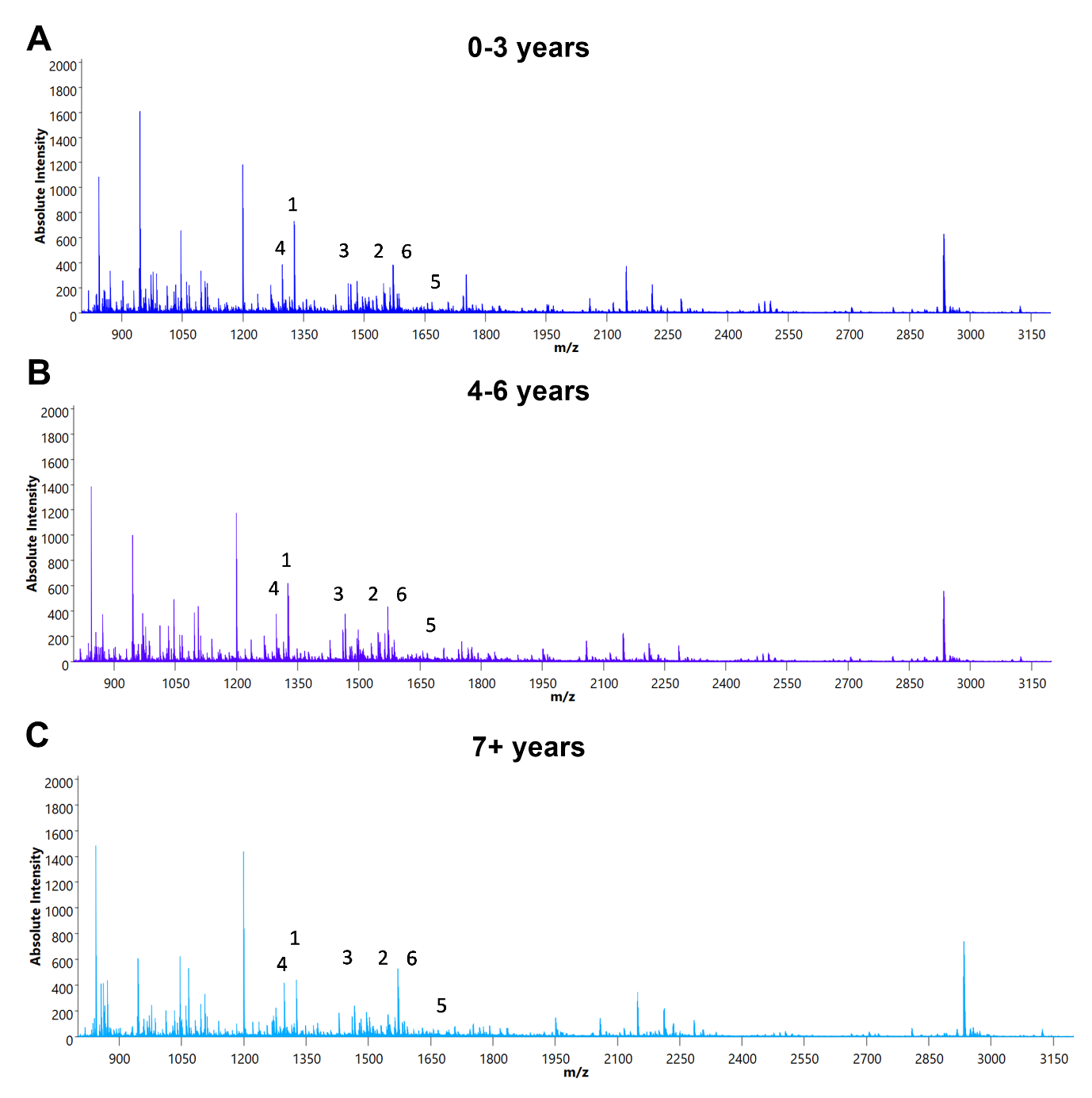

**Supplementary Figure 5.** Overall spectrum based on the three overal survival groups in stroma. (A) 0-3 years, (B) 4-6 years, and (C) 7+ years survival.

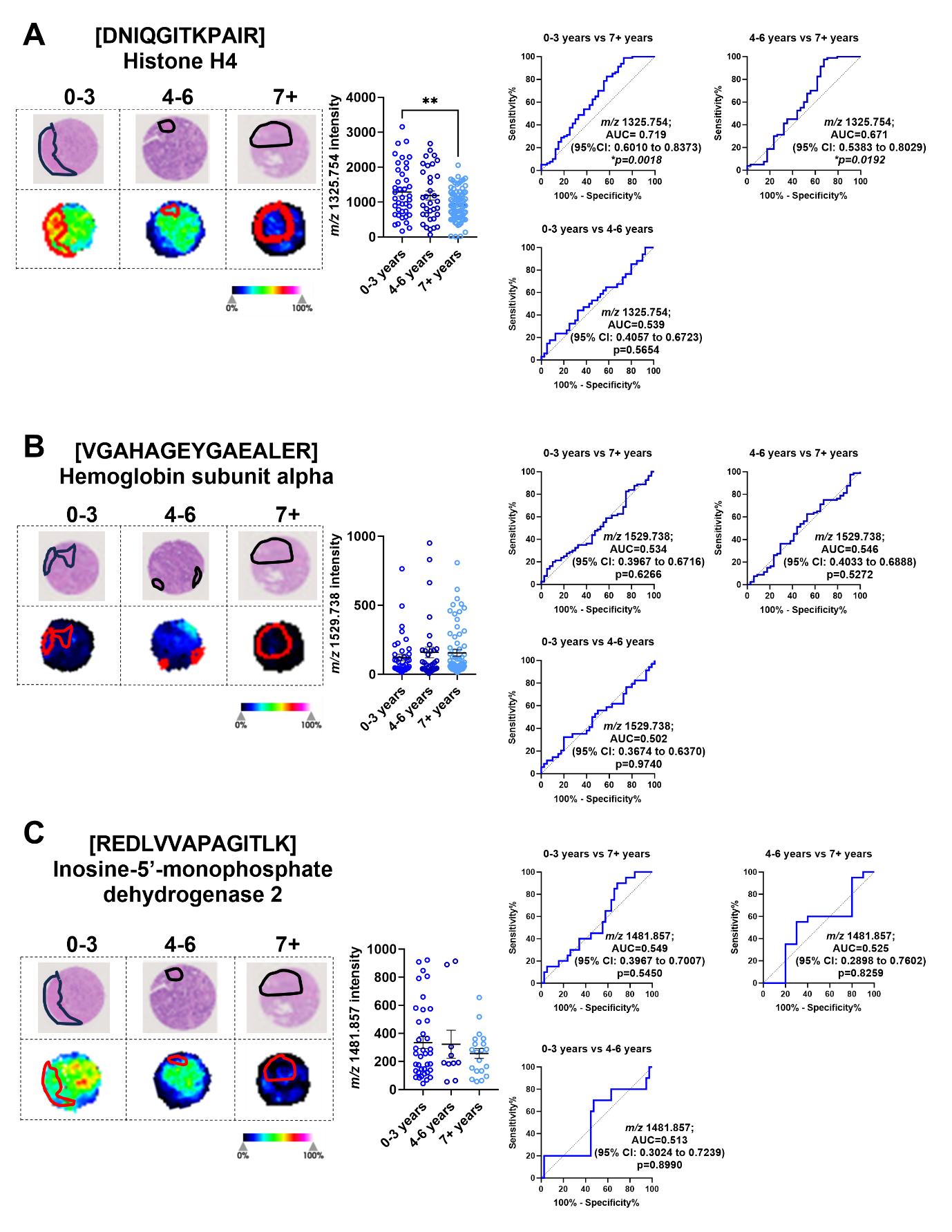

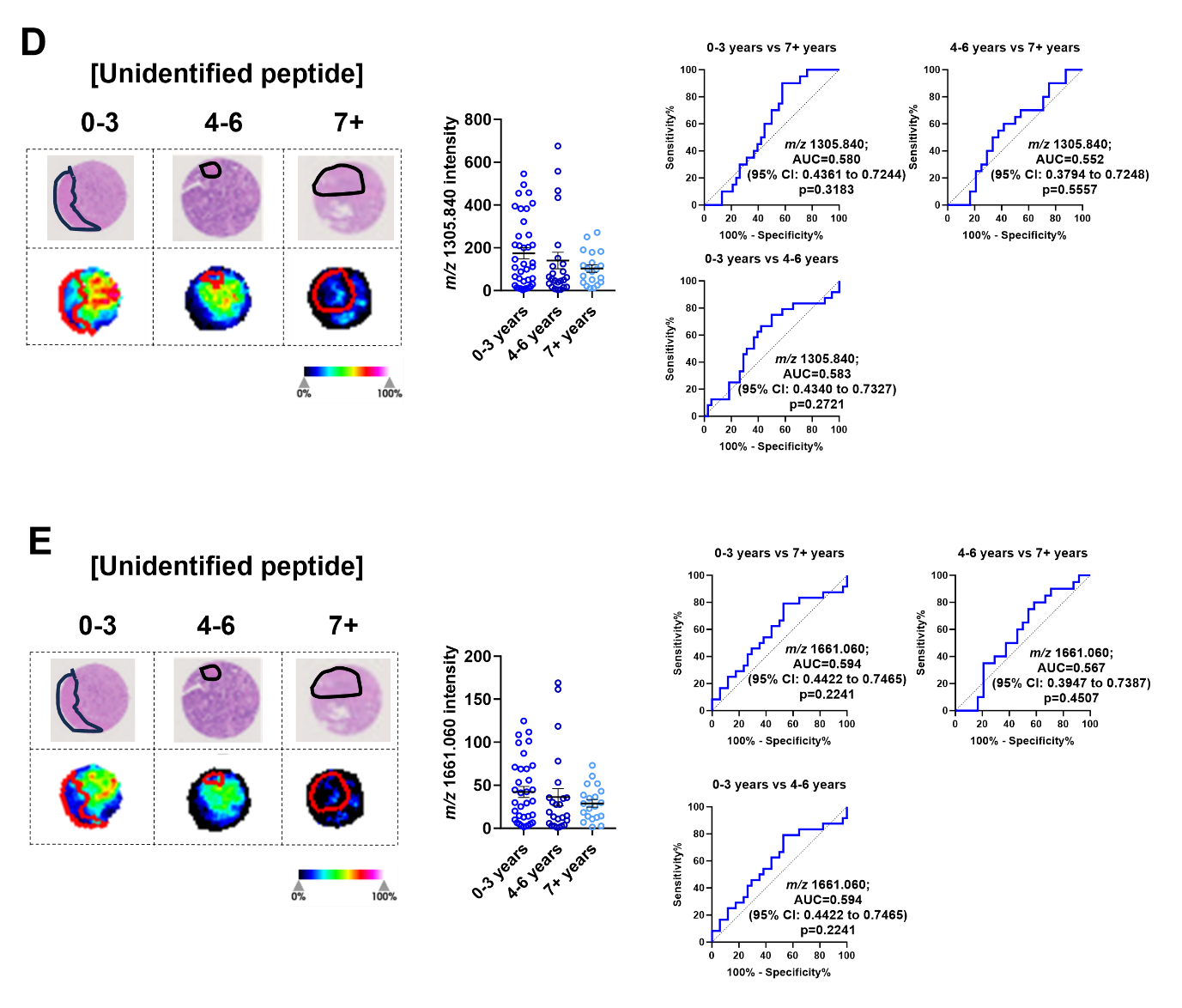

**Supplementary Figure 6.** Significantly abundant tryptic peptides associated with poor overall survival (0-3 years) in CRLM stroma regions**.** CRLM cores were subdivided based on overall survival after curative-intent surgery (0-3, 4-6, and 7+ years). Representative H&E and ion intensity maps of stroma region annotated CRLM tumour cores for *m/z* 1325.754, 1529.738, 1481.857, 1305.840 and 1661.060. Tumour cores are 1.5mm in diameter. These tryptic peptides were identified as (A) Histone H4, (B) Haemoglobin subunit alpha, (C) Inosine-5’-monophosphase dehydrogenase 2 and (D-E) unidentified by LC-MS/MS. Bar graphs showing mean ± SEM and ROC plots are presented. ***p<0.01.* Ordinary one-way ANOVA. Each dot represents a single tumour core.
